# Plasma proteomics reveals markers of metabolic stress in HIV infected children with severe acute malnutrition

**DOI:** 10.1101/19006726

**Authors:** Gerard Bryan Gonzales, James M Njunge, Bonface M Gichuki, Bijun Wen, Isabel Potani, Wieger Voskuijl, Robert H J Bandsma, James A Berkley

**Author notes:** Correspondence to: Gerard Bryan Gonzales, PhD.

## Abstract

HIV infection affects up to 30% of children presenting with severe acute malnutrition (SAM) in Africa and is associated with increased mortality. Children with SAM are treated similarly regardless of HIV status, although mechanisms of nutritional recovery in HIV and/or SAM are not well understood. We performed a secondary analysis of a clinical trial and plasma proteomics data among children with complicated SAM in Kenya and Malawi. Compared to children with SAM without HIV (n = 113), HIV-infected children (n = 54) had evidence (false discovery rate (FDR) corrected p<0.05) of metabolic stress, including enriched pathways related to inflammation and lipid metabolism. Moreover, we observed reduced plasma levels of zinc-α-2-glycoprotein, butyrylcholinesterase, and increased levels of complement C2 resembling findings in metabolic syndrome, diabetes and other non-communicable diseases. HIV was also associated (FDR corrected p<0.05) with higher plasma levels of inflammatory chemokines. Considering evidence of biomarkers of metabolic stress, it is of potential concern that our current treatment strategy for SAM regardless of HIV status involves a high-fat therapeutic diet. The results of this study suggest a need for clinical trials of therapeutic foods that meet the specific metabolic needs of children with HIV and SAM.

## Introduction

Malnutrition, specifically undernutrition in all its forms, remains a global public health burden that accounts for 45% of all death among children under 5 years old ^1^. Despite careful monitoring and adherence to guidelines set by the World Health Organization, whilst in general, uncomplicated SAM cases treated in the community do well, up to 25% of children with complicated severe acute malnutrition (SAM) treated in a hospital environment do not survive ^2–5^. Furthermore, about one in five children treated for complicated SAM and discharged alive, die in the first year after discharge in low-resource settings ^6–8^. However, our understanding of the pathophysiology underlying the poor prognosis for these children is surprisingly limited.

Infection with the human immunodeficiency virus (HIV) is a common co-morbidity of SAM in sub-Saharan Africa affecting up to 30% of admissions among SAM cases ^9^. HIV-infected or exposed children are significantly more likely to be stunted, wasted, and underweight ^10^. They also more often present with other clinical complications and greater susceptibility to infections, thus further complicating their clinical management, which may include providing more aggressive antimicrobial therapy and higher caloric nutritional intervention ^11^. Moreover, response to clinical management is also less predictable and less well-understood in HIV-infected children compared to their uninfected counterparts ^12^. Although acute opportunistic infections play a key role in the outcome of these children, intestinal pathology including inflammation and malabsorption, and metabolic perturbations may also be present. However, mechanisms driving poor nutritional recovery of children with HIV even when detected co-morbidities are treated remain poorly understood ^12^.

We hypothesised that inflammatory, metabolic and other pathways which are likely to be involved in the response to infection, survival and nutritional recovery differ between children with SAM with and without HIV. We conducted a secondary analysis of clinical data and biological samples from a randomised clinical trial in Kenya and Malawi ^13^.

## Results

### Patient characteristics

Table 1 presents the baseline characteristics of the children in the randomised trial. A total of 843 complicated SAM children were recruited for the randomised trial, of which 179 (22%) patients were HIV(+). Age was higher and MUAC was lower in HIV(+) children than HIV(-) counterparts. Most HIV cases were found in Malawi. Sex and the presence of oedema were not associated with HIV status. Mortality was more than 2 times higher among in HIV(+) compared to HIV(-) (p < 0.001). Children whose HIV status were unknown had the highest mortality of 34%, which indicates bias due to frequent death before testing could be undertaken or refusal of testing when a child was more severely ill.

**Table 1.** Descriptive characteristics of the study participants

|  | All | HIV (+) | HIV (-) | Unknown HIV status | *p* * |
| --- | --- | --- | --- | --- | --- |
| n (%) | 843 | 179 (21%) | 618 (73%) | 46 (5%) |  |
| Median age in months [IQR] | 16 [10 – 25] | 21 [12 – 31] | 16 [10 – 25] | 10 [8 – 17] | **<0.001** |
| % girls (n) | 45% (359) | 45% (81) | 45% (278) | 56% (26) | 0.95 |
| Mean MUAC in cm [95% CI] | 11.2 [11.1 - 11.3] | 10.5 [10.36 - 10.7] | 11.4 [11.3 -11.5] | 11.2 [10.9 – 11.5] | **<0.001** |
| Mean weight-for-age z-score [95% CI] | -4.01 [-4.11 – -3.92] | -4.51 [-4.72 – -4.31] | -3.92 [-4.03 – -3.80] | -3.56 [-3.94 – -3.92] | **<0.001** |
| % mortality (n) | 15% (127) | 26% (47) | 10% (64) | 34% (16) | **<0.001°** |
| % oedematous (n) | 31% (264) | 30% (54) | 33% (203) | 15% (7) | 0.50 |
| Site |  |  |  |  |  |
| Coast Provincial General Hospital, Kenya | 39% (329) | 25% (45) | 40% (247) | 80% (37) | *Reference* |
| Kilifi County Hospital, Kenya | 22% (187) | 22% (40) | 23% (145) | 4% (2) | 0.08 |
| Queen Elizabeth Central Hospital, Malawi | 39% (327) | 52% (94) | 36% (226) | 15% (7) | **<0.001** |

\*comparison between HIV(+) and HIV(-); °adjusted for age, sex and site of recruitment

Among HIV(+), 33% were already receiving an anti-retroviral treatment (ART) regime: 53/179 (30%) on highly active antiretroviral therapy (HAART), and 7/179 (4%) on Nevirapine only. About half of the children (90/179) were naïve for ART whereas HIV treatment status was unknown for 16% (29/179). Mortality was not significantly different among children on HAART, ART naïve and children with unknown HIV treatment status (Supplementary Table 1).

### HIV is associated with increased inflammatory and immune response, dysregulated lipid metabolism, and increased proteolysis in children with SAM

Among the children included in the proteomics study, 54 were HIV (+) and 113 were HIV(-) (Table 2). In this sub-population, age, sex and the presence of oedema were not significantly associated with HIV. HIV(+) children also had significantly lower MUAC and higher mortality than HIV(-) children.

**Table 2.** Patient characteristics of those subjected to proteomics analysis

|  | All | HIV (+) | HIV (-) | *p* * |
| --- | --- | --- | --- | --- |
| n | 167 | 54 | 113 |  |
| Median age in months [IQR] | 15 [10 – 26] | 15 [10 – 26] | 15 [10 – 24] | 0.433 |
| n girls (%) | 76 (45%) | 27 (50%) | 49 (43%) | 0.506 |
| Mean MUAC at admission (cm) [95% CI] | 10.9 [10.7 - 11.1] | 10.2 [9.8 - 10.5] | 11.3 [11.0 - 11.5] | **<0.001** |
| n oedematous (%) | 49 (29%) | 15 (28%) | 34 (30%) | 0.856 |
| n mortality (%) | 79 (47%) | 36 (67%) | 43 (38%) | **<0.001**<sup>°</sup> |
| *Use of antiretroviral medication* |  |  |  |  |
| Naïve |  | 27 (50%) |  |  |
| highly active antiretroviral therapy (HAART) |  | 14 (26%) |  |  |
| Nevirapine only |  | 3 (6%) |  |  |
| Unknown |  | 10 (18%) |  |  |

\*comparison between HIV(+) and HIV(-), °adjusted for age, sex, site of recruitment, oedema

A total of 204 circulating proteins were annotated and compared between children with and without HIV infection. Of these, levels of 42 proteins were found to be significantly associated with HIV status in the initial univariate analysis (Figure 1A) (Supplementary Table 2). Specifically, HIV(+) was associated with higher circulating levels of immunoglobulins, inflammatory proteins such as calprotectin (S100 calcium binding protein A8 and S100 calcium binding protein A9), complement proteins, and proteins related to host response to infection (i.e. lipopolysaccharide binding protein, galectin 3 binding protein and CD5 molecule-like protein). Enrichment analysis suggested that HIV(+) children had increased complement activation and immune response, and inflammatory responses than HIV(-) children. Neutrophil aggregation and chemokine production appeared to be the pathways most highly enriched in HIV(+) compared to HIV(-) SAM children. To substantiate these results, we quantified chemokine and cytokine levels in plasma. As shown, most chemokines had the tendency to be associated with HIV infection, where elevated plasma concentration of 12 were significantly associated with HIV status in SAM children (Figure 1B), namely: monocyte chemoattractant protein 1 (MCP1), macrophage inflammatory protein 1 beta (MIP1b, CCL4), granulocyte colony-stimulating factor (GCSF), interleukin 1 beta (IL1b), tumour necrosis factor alpha (TNFa), interleukins 2,5,7, 8 and 15 (IL2, 5, 7, 8, 15), interleukin 12 subunit beta (IL12p40), interferon gamma-induced protein 10 (IP-10), and interleukin-1 receptor antagonist (IL-1RA).

**Figure 1.**
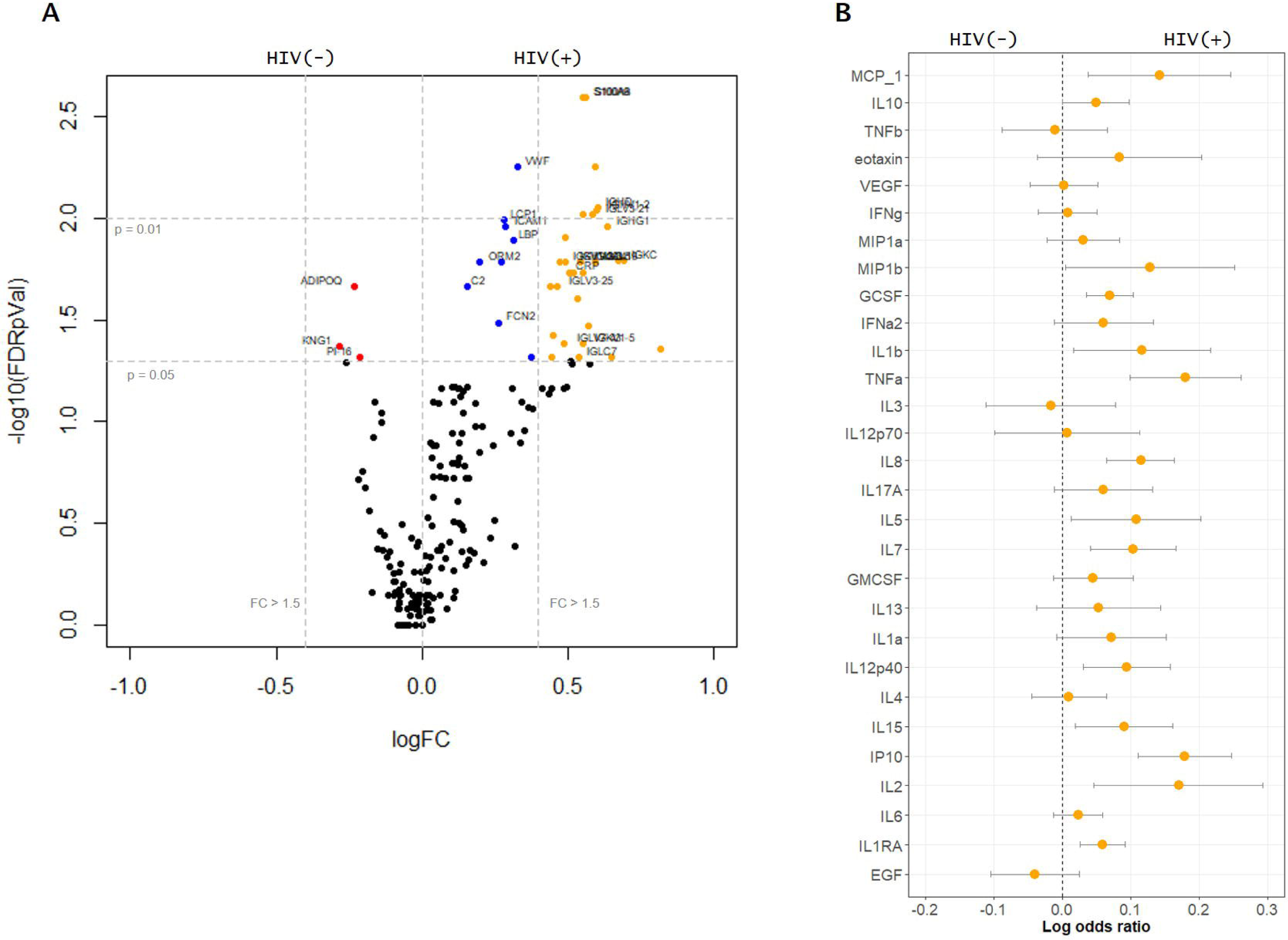
Univariate analysis of plasma proteome and individual plasma cytokines associated with HIV. (A) Volcano plot showing several significantly different (FDR adjusted *p* value < 0.05) proteins and their log2 HIV(+) versus HIV(-) fold change. Red points represent those significantly higher in plasma of HIV(-), blue points significantly enriched in plasma of HIV(+) and orange points significantly higher than 1.5 folds in HIV(+) compared to HIV(-) SAM children. Vertical lines indicate significance level at p = 0.05 and 0.01; horizontal lines indicate more than 1.5 folds enrichment. (B) Log odds plots showing association of chemokine markers analysed using Luminex platform and HIV status. Points indicate log odds ratio for every log increase in plasma protein concentration; bars indicate 95% confidence interval.

Out of the 43 differentially expressed proteins, three proteins were found to be negatively associated with HIV status on initial univariate analysis, namely: adiponectin, kininogen-1 and peptidase inhibitor 16. Among HIV(+) children, there were no statistically significant associations with receiving HAART (n = 53) compared to ART naïve (n = 90) children (data not shown), recognising our study was not powered for this comparison. Furthermore, sensitivity analysis to address the possibility of HIV maternal antibodies in younger children, showed no significant interaction of age above or below 18 months and individual proteins plasma levels towards HIV status, although power to detect was limited.

The weighted EN model extracted 73 circulating proteins (Figure 2A) that are associated with HIV status with AUROC = 0.97 [95% CI: 0.95 – 0.99] (Figure 2B) and misclassification error rate of 2.4%. Optimism-adjusted validated AUROC after bootstrapping was 0.90 [95% CI: 0.90 – 0.902], indicating a robust model. Pathway enrichment analysis highlighted that apart from increased immune response, HIV(+) children with SAM had increased proteolysis and lipid mobilisation, specifically increased very low-density lipoprotein assembly, indicating metabolic dysregulation related to cholesterol and triglyceride metabolism among HIV(+) patients (Figure 2D).

**Figure 2.**
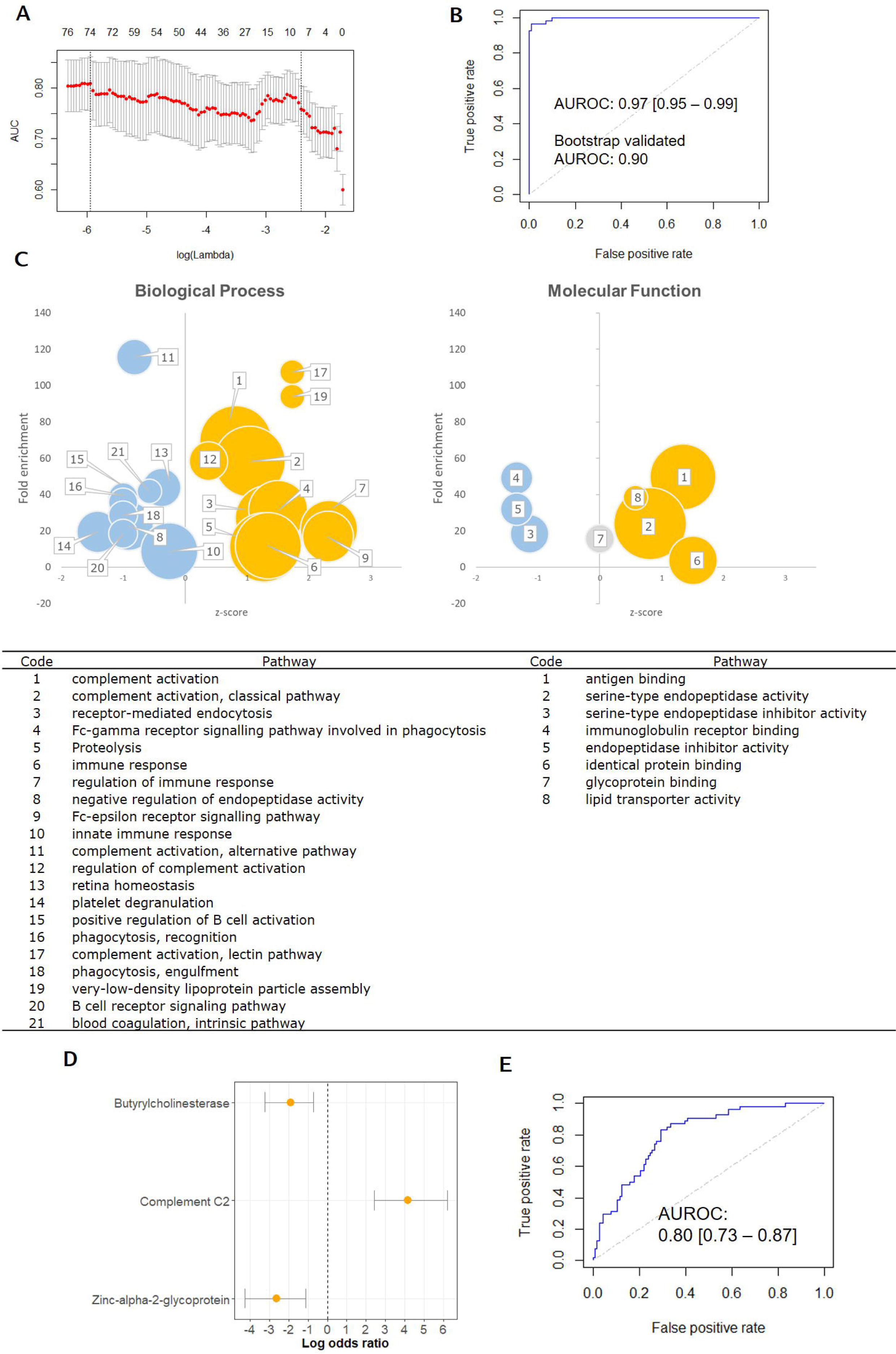
Multivariate analysis of plasma proteome associated with HIV. (A) Elastic net (EN) regularized regression lambda parameter optimization curve, optimal lambda parameter was chosen based on the highest area under the receiver operator curve (AUROC); (B) AUROC (0.97 [95% CI: 0.95 – 0.99]) of the EN model generated using the lambda parameter, alpha parameter was set to 0.75, final model extracted 34 protein features, optimism-adjusted bootstrap validation of the generated EN model, validated AUROC = 0.90 [95% CI: 0.90 – 0.90] using 2000 iterations; (C) Gene entology (GO-terms) enrichment analysis of proteins extracted by the EN model. X-axis represents z-scores; y-axis, fold enrichment, and size of the spheres represent the number of proteins involved in the particular pathway. Gold circles represent pathways enriched in HIV(+) whereas blue circles are pathways more associated with HIV(-). The grey circle indicate that there are as much proteins in this pathway that are significantly upregulated and downregulated in HIV. Only significantly enriched pathways (p < 0.05 after FDR adjustment) are plotted. See main text for explanation of the plots. Pathways enriched are identified in the table. (D) Log odds ratio plot of the 3 proteins extracted after bootstrap validation with log odds on the x-axis and bars indicating 95% confidence interval obtained using weighted logistic regression with HIV as outcome variable and the 3 proteins as covariates. Weights used were obtained by inverse probability of treatment weights; (E) Predictive ability of the weighted logistic regression model using the 3 bootstrap validated proteins with HIV as outcome variable, AUROC = 0.80 [95% CI: 0.73 – 0.87]

After 2000 bootstrap iterations during bootstrap validation, 3 proteins were consistently extracted by the EN model >80% of the time (Figure 2D), namely: butyrylcholinesterase (BChE), complement C2 and zinc-α-2-glycoprotein (ZAG), indicating that these 3 proteins are likely to be the most important features associated with HIV in children with complicated SAM. Weighted logistic regression model of these 3 proteins showed good discrimination of HIV status (AUROC = 0.80 [95% CI: 0.74 – 0.87]) (Figure 2E).

## Discussion

In this study, we report plasma proteomic differences associated with HIV status, suggesting that HIV imposes additional metabolic and inflammatory insults among HIV(+) children with SAM. Our results show that pathways involved in inflammatory response, complement cascade activation and lipid metabolism dysregulation are associated with HIV status. Circulating levels of several plasma chemokines were also found to be higher in HIV(+) among children with SAM. Greater inflammatory responses in these children could be related to the higher inpatient mortality of HIV(+) compared to HIV(-) children with SAM.

An earlier metabolomics study in Uganda reported reduced serum levels of adiponectin and leptin, whereas serum triglycerides, ketones and even-chain acylcarnitines were higher in HIV(+) children with SAM indicating perturbed lipid metabolism ^14^. Our current study therefore concurs with this finding, as we also found reduced plasma levels of adiponectin in HIV(+) SAM children compared to HIV(-) SAM children, along with upregulation of pathways involved in lipid transport and metabolism, specifically very low-density lipoprotein assembly.

Using optimism-adjusted bootstrap validation of the EN model, we found 3 proteins: complement c2, BChE and ZAG robustly distinguished HIV(+) from HIV(-) in children with SAM, demonstrating the ability of multivariate analysis techniques, such as EN, to uncover underlying relationships between protein markers which would be difficult to identify when analysed individually. The activation of the complement system during HIV infection has been previously discussed at length, which is associated with the increased cellular invasion of HIV in cells ^15–17^.

On the other hand, BChE is a protein synthesized in the liver and abundant in plasma, which hydrolyses acetylcholine. Although very similar to its sister protein, acetylcholinesterase, biological functions of BChE appear to be more varied but less understood ^18^. In a recent study in China, low circulating BChE was found to be highly associated with HIV severity, was predictive of mortality in adults, and was proposed as a plausible strategy for severity classification among adults with HIV ^19^. BChE is also reported to be reduced in SAM, stress and inflammation ^20^. In animal studies, BChE deficiency was found to strongly affect fat metabolism and promotes hepatic lipid accumulation ^21^. Serum BChE levels have been found to have a significant negative correlation with serum total cholesterol and serum low-density-lipoprotein cholesterol among people with diabetes mellitus ^22^.

ZAG is a newly characterized adipokine that is involved in lipolysis, body weight regulation and may also be involved in the development of insulin resistance ^23^. Reduction in plasma levels of ZAG was previously reported to be implicated in dyslipidaemia in HIV(+) adults under ART treatment ^23^. Reduced circulating levels of ZAG has also been found among adults with clinically diagnosed metabolic syndrome, based on guidelines of the United States National Cholesterol Education Program (NCEP) Expert Panel Adult Treatment Panel (ATP) III criteria ^24^. Serum ZAG levels have been reported lower among adults with impaired glucose tolerance and type 2 diabetes mellitus ^25^. Taken together, our results therefore suggest that children with both HIV and SAM manifest hallmarks of metabolic stress similar to those occurring in metabolic syndrome and other non-communicable diseases (NCD).

This study is the first proteomics investigation on the interaction between HIV and SAM. In summary, our results, which together with the previously published metabolomics study ^14^, strengthens evidence on the increased metabolic stress and altered metabolic response among children living with both HIV and SAM. Our results also concur with previous studies that reported elevated metabolic stress among non-malnourished adults living with HIV leading to increased prevalence or risk for metabolic syndrome, cardiovascular diseases, diabetes and other non-communicable diseases ^26–32^.

Metabolic abnormalities have previously been reported to be attributed HAART use among HIV(+) patients ^33^. In a recent systematic review, use of 2 classes of HAART, protease inhibitors and nonnucleoside reverse transcriptase inhibitors, has been found to be associated with abnormalities in plasma lipid profiles ^34^. However, dysregulation in lipid metabolism has also been reported in HAART-naïve patients, which indicates that HIV infection alone cause lipid metabolism perturbations. An earlier longitudinal study of 50 men in the USA reported notable declines in serum total cholesterol after HIV infection compared to results of blood analysis from last seronegative visit. Large increases in total cholesterol and low-density lipoproteins (LDL) were detected after HAART initiation ^35^. However, many other studies reported increases in total cholesterol among HIV-infected patients naïve to HAART. For instance, in a study of ART-naïve HIV-infected adults in Ethiopia, malnutrition and lipid abnormalities (specifically total cholesterol) were associated with CD4+ T cell counts ^36^. In in vitro studies, transfection of a T-cell (RH9) with HIV led to the enhanced production of free fatty acids and LDL ^37^. Furthermore, monocytes isolated from HIV-infected patients both taking HAART and HAART-naïve, were found to have altered expression patters of receptors linked with lipid metabolism (i.e. FXR, PXR, PPARα, GR, RARα and RXR) compared to monocytes of HIV-uninfected controls ^38^. For our study however, we are unable to ascertain whether the lipid metabolism dysregulation we observed is due primarily on the viral load itself or the use of HAART due to lack of power for this sub-analysis. Majority of the participants subjected to proteomics analysis were HAART-naïve (50%), where 26% were on HAART, 6% were on Nevirapine alone and we had no data on treatment of 18% of the patients (Table 2). In all these studies cited, authors argue to need for monitoring of lipid profiles in HIV-infected populations. Hence, lipid monitoring may also inform nutritional and clinical recovery of children with SAM and HIV and could be implemented to improve clinical care for these children.

However, despite our knowledge that HIV-infected populations have altered metabolic requirements compared to HIV-uninfected counterparts, WHO guidelines for the nutritional management for SAM are globally the same regardless of HIV status, which is summarized in Table 3 ^39^. Nutritional management for in-patient children with SAM involves provision of a low-protein, low-fat milk-based food, F75, every three hours. F75 is used during clinical stabilization occurring during the first few days after admission and is not intended for weight gain. Once the children are clinically stabilized and are able to tolerate the milk/solute load, children are transitioned to F100, a higher-calorie, high-fat milk intended to boost weight gain or to Ready-to-Use Therapeutic Food (RUTF), a peanut-based calorie-dense diet. Upon discharge from in-patient care, children are referred to community based nutritional therapeutic centres where they are provided with RUTF on a 2 weekly basis.

**Table 3.**
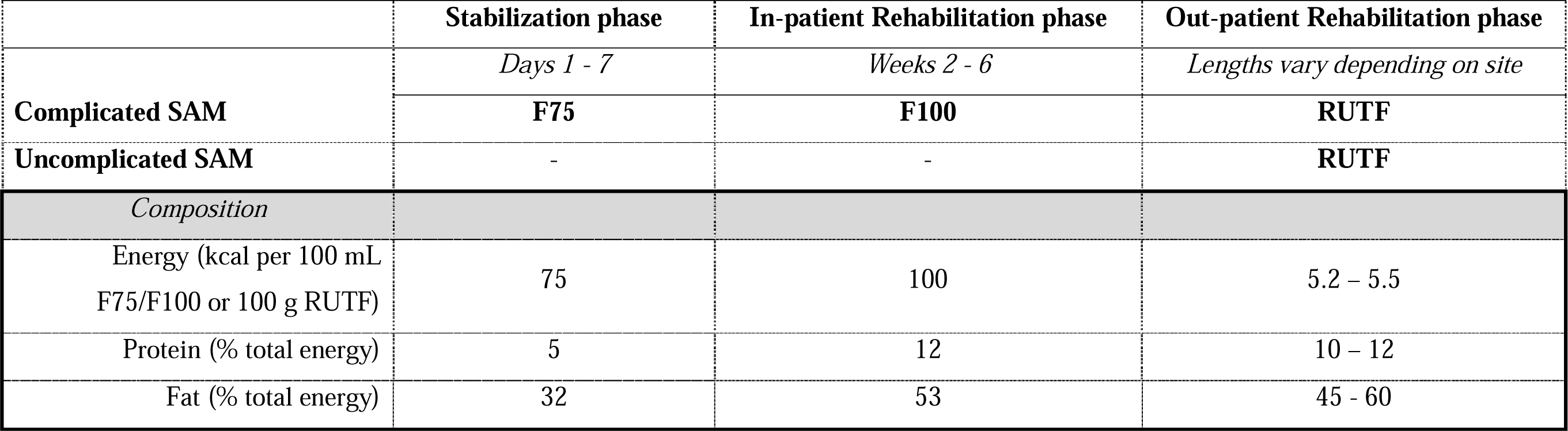
Nutritional management protocol for children with severe acute malnutrition ^39^

|  | **Stabilization phase** | **In-patient Rehabilitation phase** | **Out-patient Rehabilitation phase** |
| --- | --- | --- | --- |
|  | *Days 1 - 7* | *Weeks 2 - 6* | *Lengths vary depending on site* |
| **Complicated SAM** | **F75** | **F100** | **RUTF** |
| **Uncomplicated SAM** | - | - | **RUTF** |
| *Composition* |  |  |  |
| Energy (kcal per 100 mL<br>F75/F100 or 100 g RUTF) | 75 | 100 | 5.2 – 5.5 |
| Protein (% total energy) | 5 | 12 | 10 – 12 |
| Fat (% total energy) | 32 | 53 | 45 - 60 |

Considering evidence of biomarkers of metabolic syndrome and NCD in HIV(+) children with SAM, it is of potential concern that our current treatment strategy involves a high-fat therapeutic diet. About 50% of much needed calories during the growth catch-up phase are supplied as lipids, which HIV(+) children may not be able to efficiently assimilate. Alterations in lipid metabolism in HIV(+) children with SAM may also mean that the high amounts of dietary lipids could be deposited as ectopic fat in the liver and muscle, predisposing to insulin resistance, diabetes, cardiovascular problems and other NCDs later in life. Although long-term metabolic follow-up studies could be done for HIV(+) children previously treated for either complicated and uncomplicated SAM, significant barriers are the high mortality rate in earlier studies of HIV(+) children with SAM, cost and difficulty tracing them years later. The results of this study indicate a need for clinical trials of F100 or RUTF modified to meet the expected metabolic needs of HIV(+) children with SAM. This could initially be done in relatively small groups with outcomes that include measuring metabolic stress.

Several studies on nutritional intervention strategies among HIV-infected adults have been reported. For instance, a study in the USA showed that dietary fat intake, specifically saturated fats, was significantly associated with hypertriglyceridemia among HIV-infected adults (18 – 60 years) ^40^. Moreover, in a preclinical model, high saturated fat consumption was found to accelerate immunodeficiency virus disease progression in macaques, specifically increased mortality hazard and circulating levels of pro-inflammatory cytokines, especially IL8 ^41^, which has been previously reported to be associated with lipodystrophy among HIV patients ^42^. In our study, we also found a significant association between high plasma IL8 concentration and HIV in SAM children. Hence, modifying the saturated fat composition of the milk-based F75 and F100 could potentially lower metabolic stress.

The European Society for Parenteral and Enteral Nutrition (ESPEN) have given a grade A recommendation for the use of medium-chain triglyceride (MCT)-based diet on HIV(+) patients with diarrhoea and severe undernutrition in its 2006 ESPEN Guidelines on Enteral Nutrition ^43^. Grade A recommendations are given to strategies based on meta-analysis or at least one randomised control trial. In this case, the recommendation was based on a prospective, randomized double-blind comparative trial on 24 adult patients with HIV and diarrhoea of more than 4-wk duration, fat malabsorption, and loss of 10–20% of ideal body weight ^44^. In this study, the authors found improved outcomes from diarrhoea and fat malabsorption from MCT than long-chain triglyceride-based diet among HIV(+) adults.

Lastly, the long-term metabolic effect of nutritional intervention strategies for SAM still remains unresolved. Most specifically, the potential metabolic stress associated with the rapid weight gain during the nutritional rehabilitation phase after SAM and its implications on nutritional outcomes during adulthood demands urgent research attention, especially for HIV(+) children with SAM.

Limitations of this study include absence of data on viral load and CD4+ counts of the patients, which could provide a deeper understanding of the results. Furthermore, in this study, we did not find association between oedematous malnutrition and HIV status, although several studies have a found higher HIV prevalence among non-oedematous children with SAM ^45–47^. In our study however, we found high in-patient mortality rate (16/46, 34%) among children with unknown HIV status, where 39/46 (85%) had non-oedematous SAM. Considering the high rate of mortality, these children may have been HIV(+). This highlights the need for earlier HIV screening among children with SAM.

### Conclusion

Plasma proteomics reveals that HIV(+) children with SAM manifest hallmarks of metabolic stress similar to those observed in non-communicable diseases. This could be related to the poor nutritional recovery and high mortality of HIV(+) children with SAM despite clinical and nutritional intervention. The results of this study indicate a need for clinical trials modifying the composition of F100 or RUTF to meet the specific metabolic needs of HIV(+) children with SAM during rehabilitation phase. This could initially be done in relatively small groups with outcomes that include measuring metabolic stress.

## Methods

### Patient recruitment and study design

This is a secondary analysis of a nested case control study from a randomised controlled trial (NCT02246296), which tested the effect of a lactose-free, low-carbohydrate F75 milk to limit carbohydrate malabsorption, diarrhoea and refeeding syndrome among children hospitalized for complicated SAM at Queen Elizabeth Central Hospital in Blantyre, Malawi, Kilifi County Hospital and Coast General Hospital, Mombasa, Kenya ^13^. Children aged 6 months to 13 years were eligible for enrolment into the trial at admission to hospital if they had SAM, defined as: mid-upper arm circumference (MUAC) <11.5cm or weight-for-height Z score <−3 if younger than 5 years of age, BMI Z score <−3 if older than 5 years, or oedematous malnutrition at any age and had medical complications or failing an appetite test, as defined by WHO guidelines ^48^. Children were excluded if they had a known allergy to milk products and did not provide consent. Biological samples were obtained before the children received the randomised treatment irrespective of HIV status. Unless a child’s HIV positive status was documented, HIV status was assessed by offering an antibody test at admission plus appropriate counseling. For this analysis, patients that tested positive on an HIV antibody test were considered HIV(+) and children with missing or declined HIV test were excluded.

The nested case-control study was designed to investigate inpatient mortality for which proteomic, cytokine, and chemokine data was generated using plasma samples collected at admission during enrolment to the trial. To compare the proteomic profiles between HIV affected and non-affected children with SAM, we used data from a nested case-control study to investigate inpatient mortality during the trial including 79 deaths and 88 children that were clinically stabilized and discharged from the hospital matched by site of recruitment. For this analysis, all observations from the proteomics dataset were included and the analysis was designed to help overcome selection bias, as given in detail below.

### Proteomics, cytokine and chemokine analysis

Untargeted proteomics and targeted cytokines and chemokines analysis of plasma samples were performed following methods described previously ^49^. The targeted protein panel included: epidermal growth factor (EGF); eotaxin; granulocyte-colony stimulating factor (GCSF); granulocyte-macrophage colony-stimulating factor (GMCSF); interferon alpha-2 (IFNa2); interferon gamma (IFNg); interleukins 10, 12p40, 12p70, 13, 15, 17A, 1a, 1b, 1RA, 2 to 8; interferon gamma-induced protein 10 (IP10); monocyte chemoattractant protein 1 (MCP1), macrophage inflammatory protein 1 alpha and beta (MIP1a & b); tumour necrosis factor alpha (TNFa) and beta (TNFb); and vascular endothelial growth factor (VEGF).

### Data analysis

Data analyses were performed using R v3.5 ^50^. Analysis of the prevalence of HIV(+), nutritional status and their associations with inpatient mortality utilised the entire trial dataset (N=843). Analysis of categorical data was performed using Fisher’s test and generalised linear models for continuous outcomes. Logistic regression was used to analyse binary outcomes adjusting for age, sex, presence of oedema, and site of recruitment. These associations were also adjusted for MUAC. As a sensitivity analysis to address the possibility of confounding due to HIV maternal antibodies in younger children, a test of interaction between age above or below 18 months and individual proteins towards HIV status was performed.

The proteomics, cytokines and chemokines analyses were secondary analyses of data collected from a nested case-control study with inpatient mortality as its primary outcome, hence with strong selection bias. The analysis for the association between HIV status and individual proteins was therefore performed using logistic regression analysis with inverse probability weighting (IPW) to correct for selection bias ^51–54^. Weights (*w*) were calculated as suggested by Samuelsen ^52^ wherein the weight for each observation selected into the nested case-control study was computed as the inverse of the probability of being selected for the nested study from the main clinical trial. The probability of inclusion was therefore calculated as:

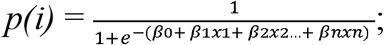

where *p(i)* is the probability of inclusion in the nested case-control study and x1, x2 … xn are HIV status, sex, age, presence of oedema, mid-upper arm circumference, and site of recruitment of the *i-*th observation (child) based on the entire trial population. Inverse probability weight is therefore:

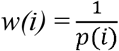

Differences in individual proteins abundances were considered statistically significant when *p*<0.05 after adjustment for multiple comparisons using Benjamini-Hochberg false discovery rate (FDR) ^55^.

Multivariate analysis was undertaken in order to determine several proteins that are collectively associated with HIV status, some of which may not be significantly associated to HIV independently. This was performed using a weighted elastic net (EN) model implemented using the “glmnet” package in R ^56^. EN is a penalized regression approach that was developed to help overcome problems caused by high dimensional data. It is an integration of two regularized approaches, ridge regression and least absolute shrinkage and selection operator (LASSO), wherein the contribution of each of these models to the final EN model is controlled by the α parameter ^56,57^. The strong penalization imposed by LASSO draws coefficients to zero thereby eliminating non-predictive proteins features, whereas ridge regression addresses potential multi-collinearity problems in high-dimensional data ^56,57^.

Weighted EN model generation was performed with HIV status as outcome, protein profile as predictors, and *w* as observation weights. The penalization parameter lambda, which influences the shrinkage of variable coefficients to zero thus eliminating some non-contributing variables, was determined by estimating the area under the receiver operator curve (ROC) of the population using ten-fold cross validation. Several alpha parameter values were assessed and a final value of 0.85 was taken to achieve a compromise between predictive ability and fewer number of features extracted. The final lambda parameter was based on the value which gave the highest area under the ROC (AUROC) value.

Proteins with significant association with HIV status after correction for false discovery and those extracted by the EN model were then uploaded to The Database for Annotation, Visualization and Integrated Discovery (DAVID) v6.8 Bioinformatics Resource ^58^ to assess the Gene ontology (GO) enriched pathways of the differentially expressed proteins.

EN model validity was judged based on the AUROC and misclassification error rate. The fitted EN model performance measured as optimism-corrected AUC was validated using bootstrap, following the procedure of Smith et al. ^59^. Bootstrapping was performed on 2000 iterations using the “BootValidation” package in R. Protein features extracted at least 80% of all iterations by the bootstrap EN model were then considered to be the most relevant protein biomarkers. To test how well these proteins can discriminate HIV status, they were then fitted on a weighted logistic regression with HIV as outcome.

### Visualisation of significantly enriched GO terms

Bubble plots were used to visualise the significantly enriched pathways (p<0.05 after adjustment for FDR) obtained from DAVID. The p-values in DAVID were obtained using a modified Fisher’s exact test ^60^. The y-axis represents the fold enrichment which indicates the magnitude of the enrichment, as calculated in DAVID. Fold enrichment is defined as:

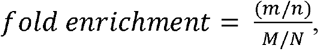

where *m* is the number of proteins significantly associated with HIV status or proteins extracted by the EN model that belong to a particular pathway, while *M* is the total number of proteins belonging to the same pathway. Variable *n* is the number of all proteins significantly associated with HIV status or extracted by the EN model and *N* is the total number of all proteins in the human background. Therefore, a fold enrichment of 10 indicates that 10% of the proteins significantly associated with HIV status belong to a particular pathway, and 1% of all annotated proteins in the human background belongs to the same pathway ^60^. However, the proponents of this metric warn that big fold enrichments could be obtained from a small number of proteins, which could be due to small *n* or pathways with fewer members.

The x-axis on the hand represents the enrichment z-score for a particular pathway ^61^, which is calculated as follows:

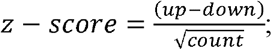

where *up* is the total number of proteins upregulated, *down* is the total number of proteins downregulated, and *count* is the total number of proteins in the input which belongs to a particular pathway. Variables *up* and *down* were based on the weighted logistic regression for each individual protein. Hence, if 5 proteins belonging to pathway *x* were upregulated and 2 were downregulated, the z-score for pathway *x* would be: (5-2)/√7 = 1.13. A positive z-score indicates that the particular pathway is overall upregulated in HIV(+), whereas a negative z-score indicates an overall downregulation ^61^.

## Data Availability

Request for data access can be sent to KEMRI-Wellcome Trust via Prof James Berkley. Data is currently not available in public repositories but the authors are currently in the process of obtaining permission to be able to make it publicly available.

## Study approvals

The secondary analyses of the trial were approved by the Kenyan national ethics committee, KEMRI-SERU (KEMRI/RES/7/3/1). The trial was registered at clinicaltrials.gov (NCT02246296).

## Author contributions

GBG, JMN and JAB designed the study and data analysis. JMN and BG performed the proteomics analysis. GBG performed data analysis. BW, RB, WV, IP and JAB were involved in the clinical aspects of the study. GBG wrote the initial drafts of the manuscript and all authors contributed to editing and improving the manuscript.

## Tables

**Supplementary Table 1.** Prior use of anti-retroviral treatment among all HIV (+) children included in the clinical trial

|  | **Prior HIV treatment\*** |  |  |  |
| --- | --- | --- | --- | --- |
|  | **HAART** | **NVP only** | **ART naïve** | **Unknown** |
| n (%)° | 53 (30%) | 7 (4%) | 90 (50%) | 29 (16%) |
| Mean MUAC at admission (cm) [95% CI] | 10.9 [10.5 - 11.4] | 9.8 [8.9 - 10.6] | 10.5 [10.2 - 10.8] | 9.6 [8.1 - 11.1] |
| Mortality n (%) | 10 (18%) | 1 (28%) | 25 (28%) | 1 (25%) |
| Odds ratio for mortality [95% CI] | 0.97 [0.82 - 1.12]<br>p=0.63<sup>#</sup> | - | *Reference* | 1.07 [0.88 - 1.30]<br>p=0.45<sup>#</sup> |

\*HAART - highly active antiretroviral therapy; NVP – Nevirapine; <sup>#</sup> adjusted for age, sex, site and oedema

**Supplementary table 2.**
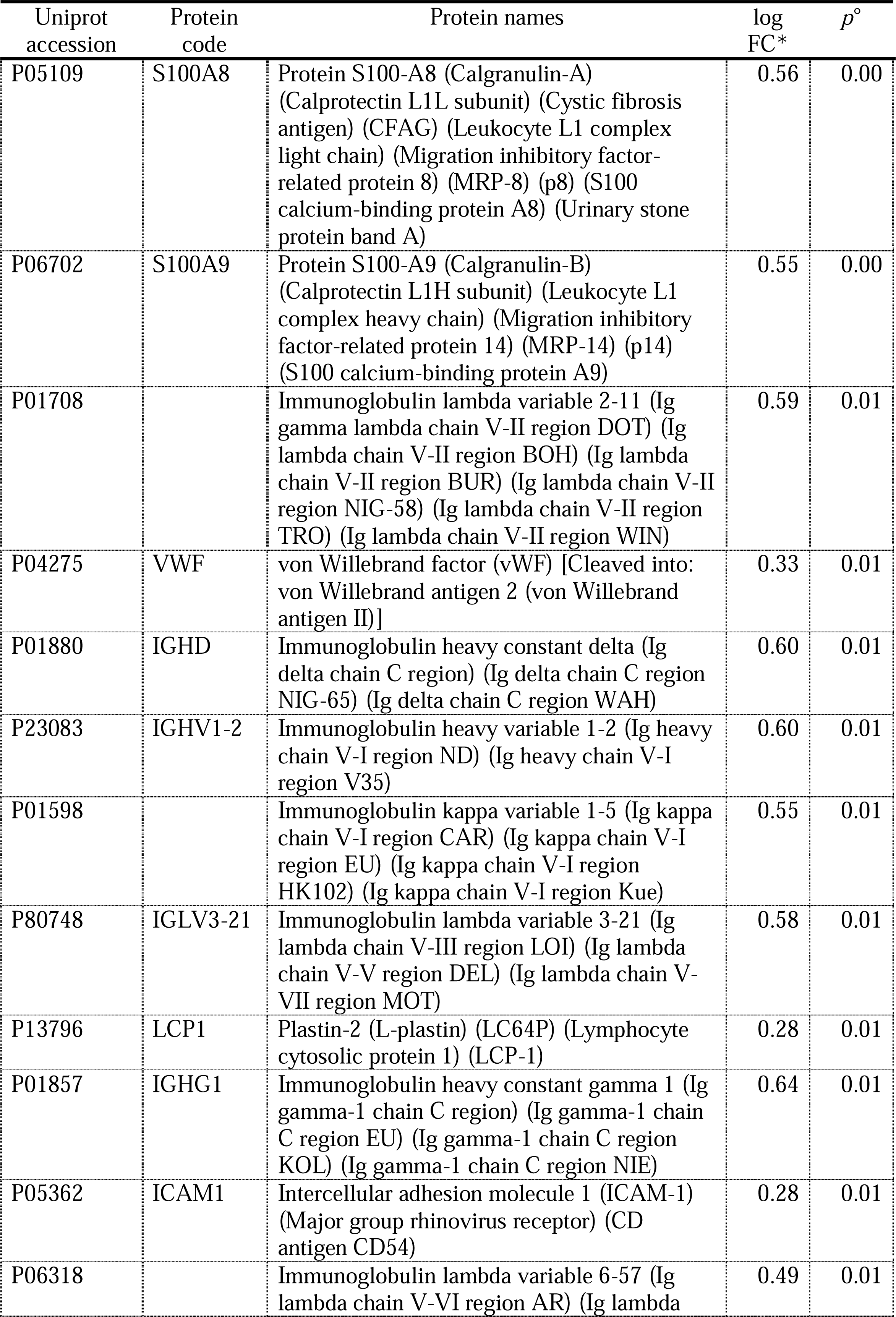
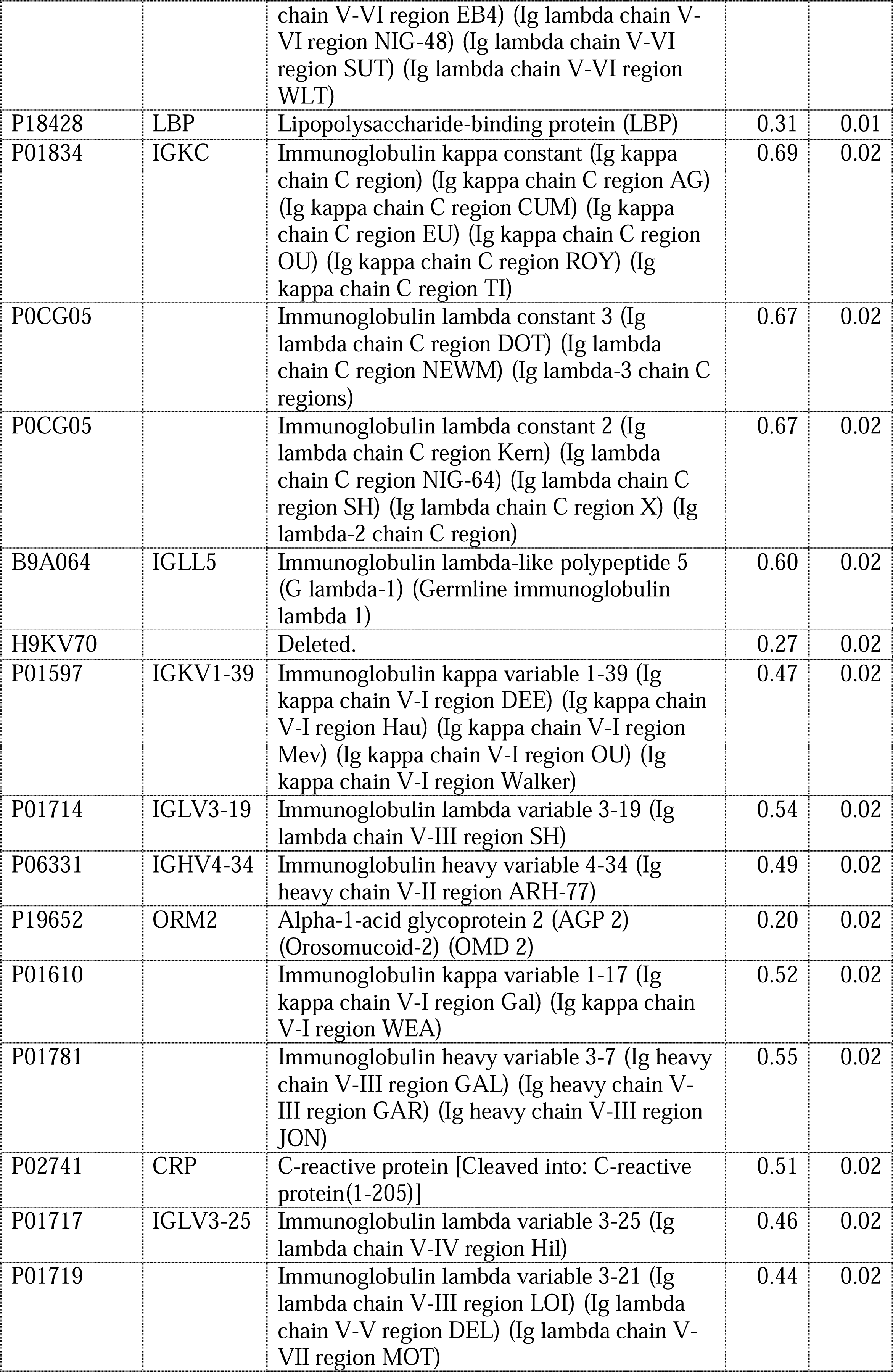
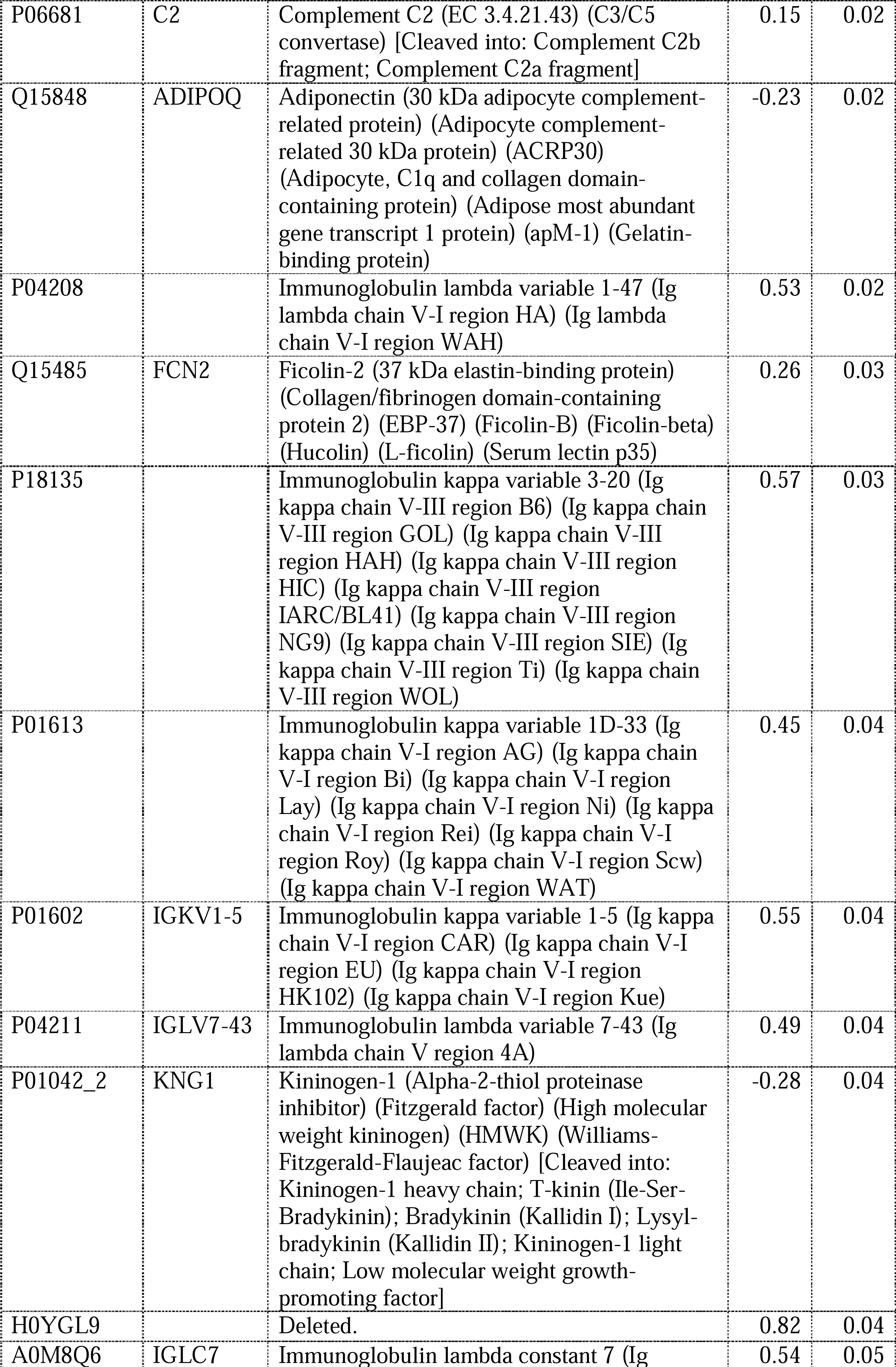
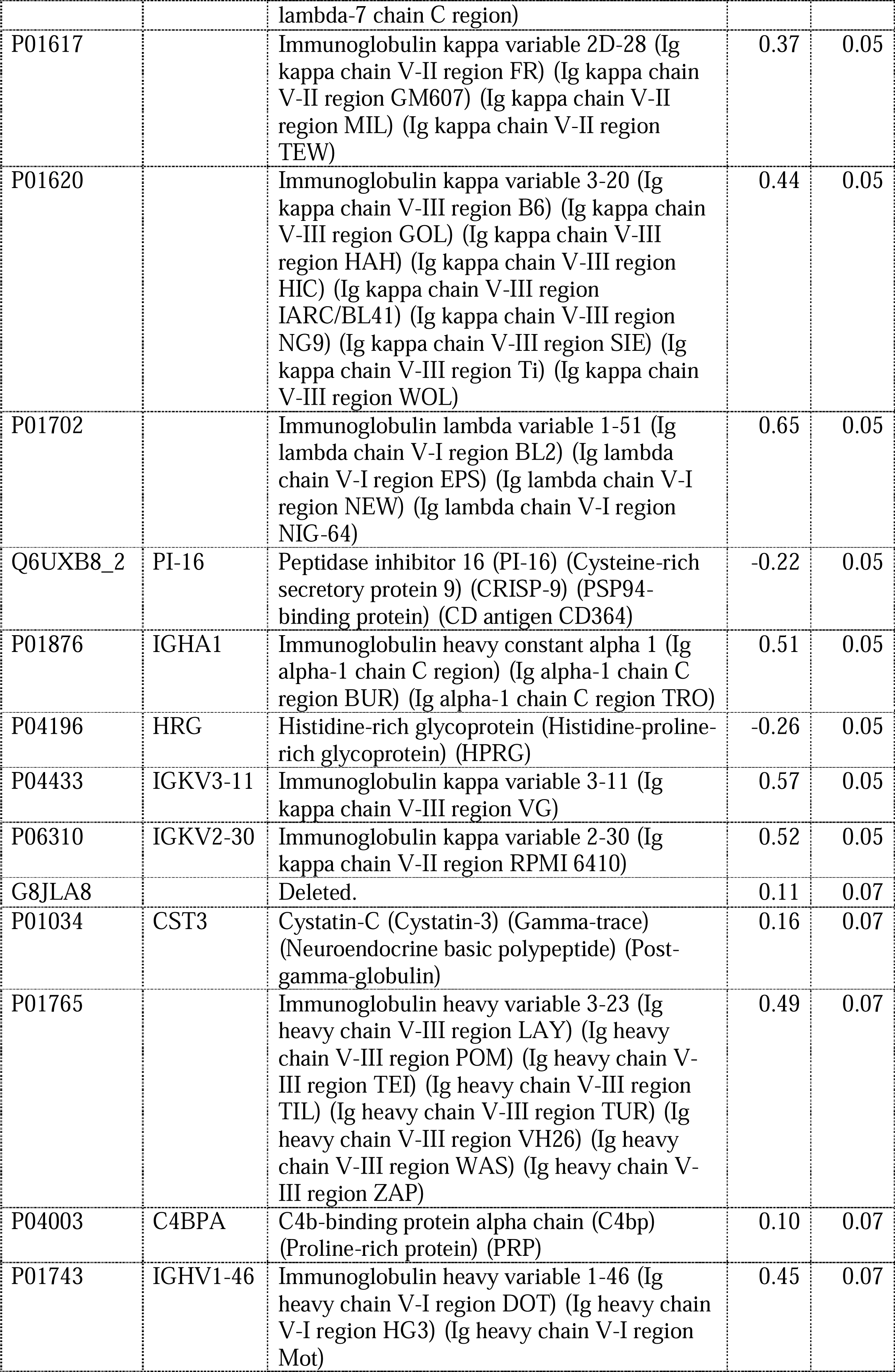
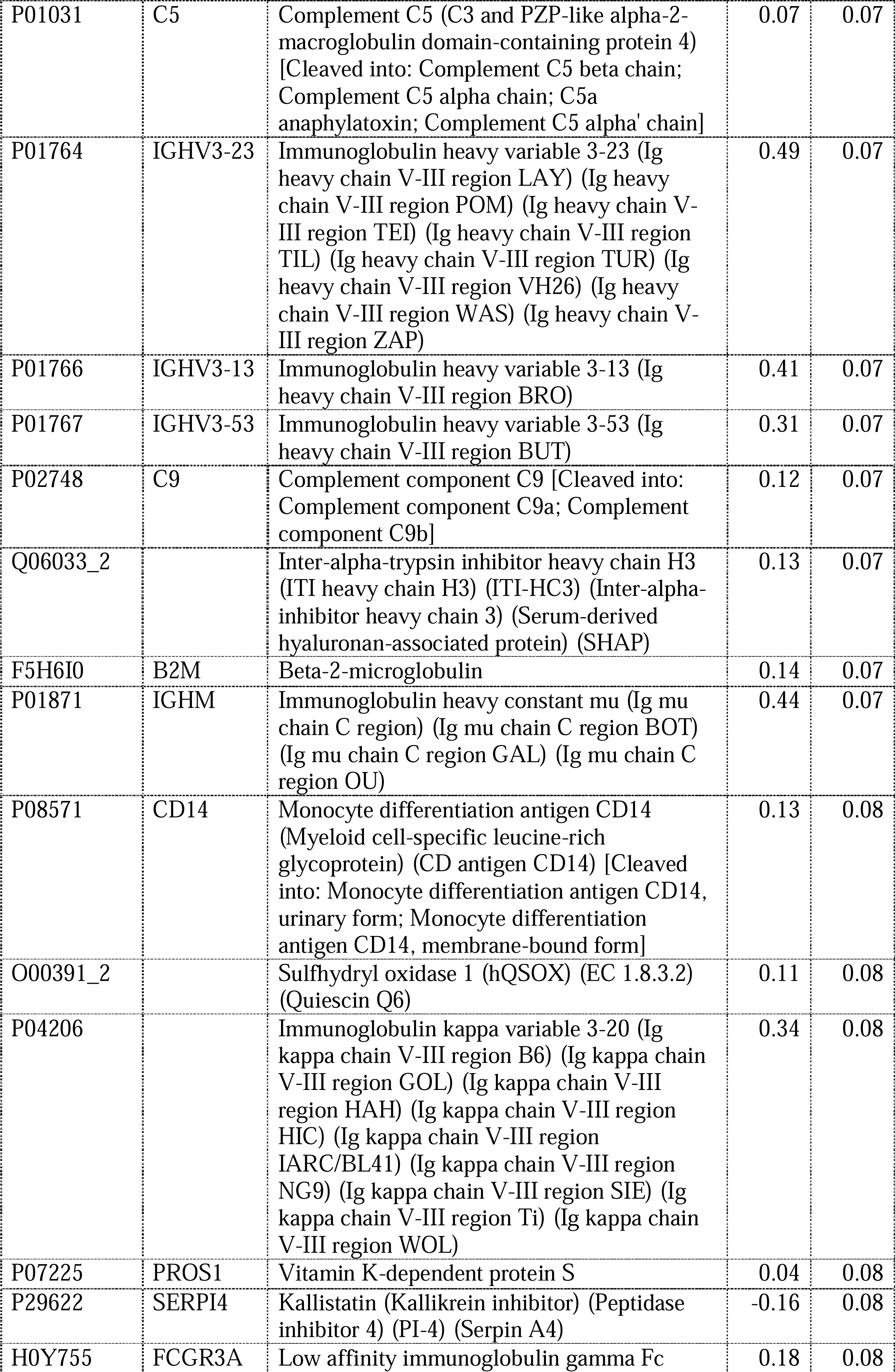
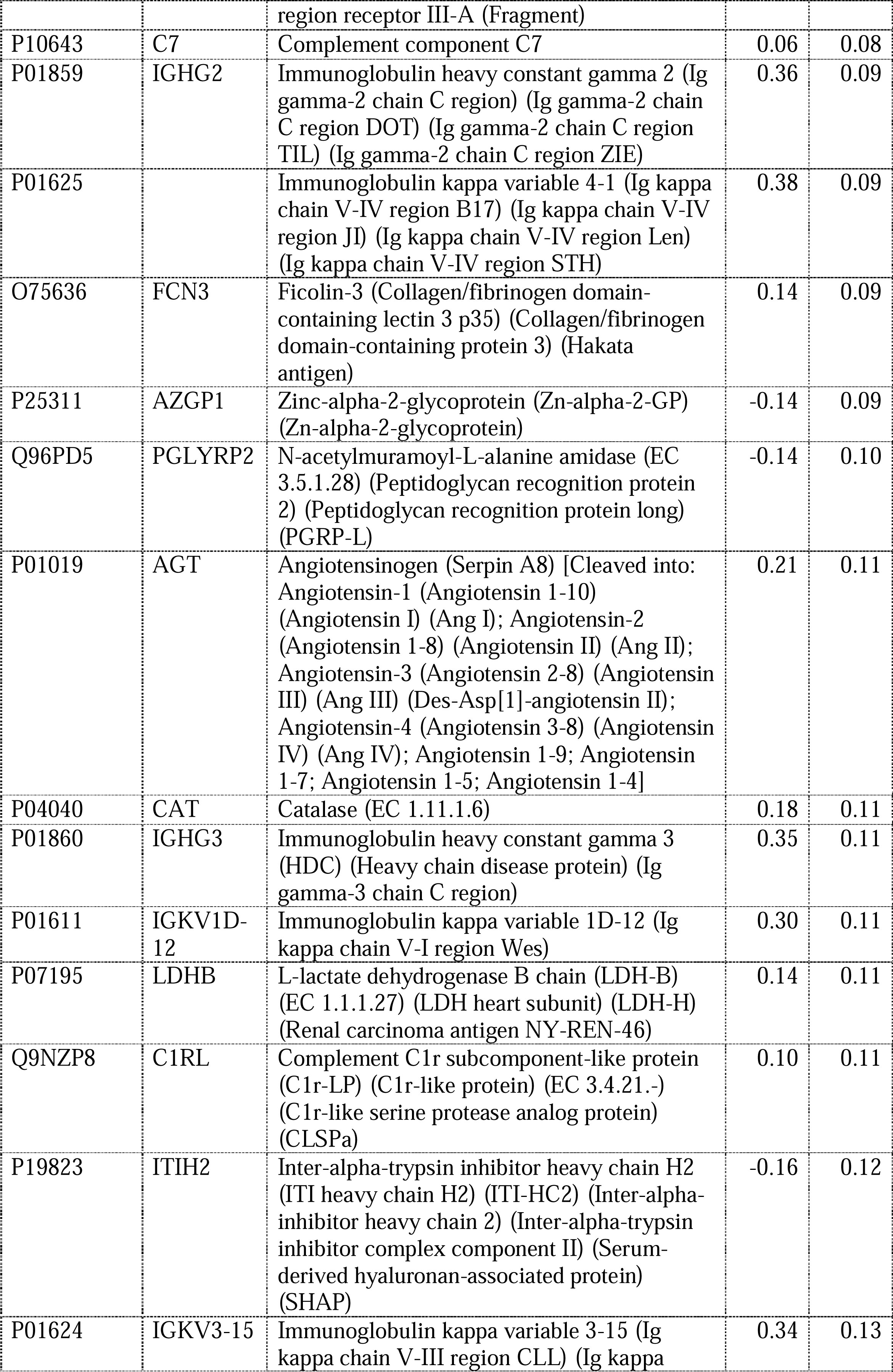
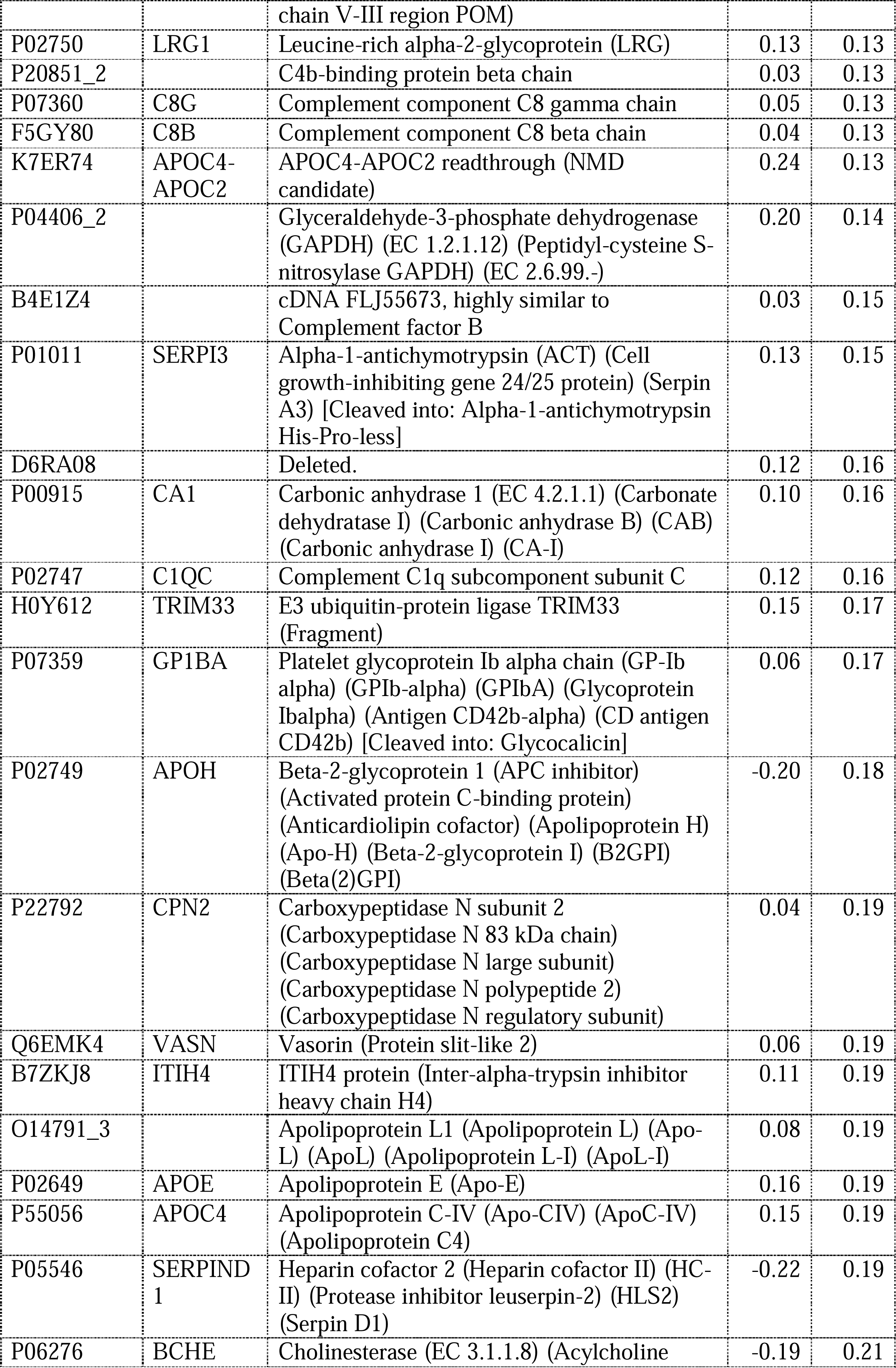
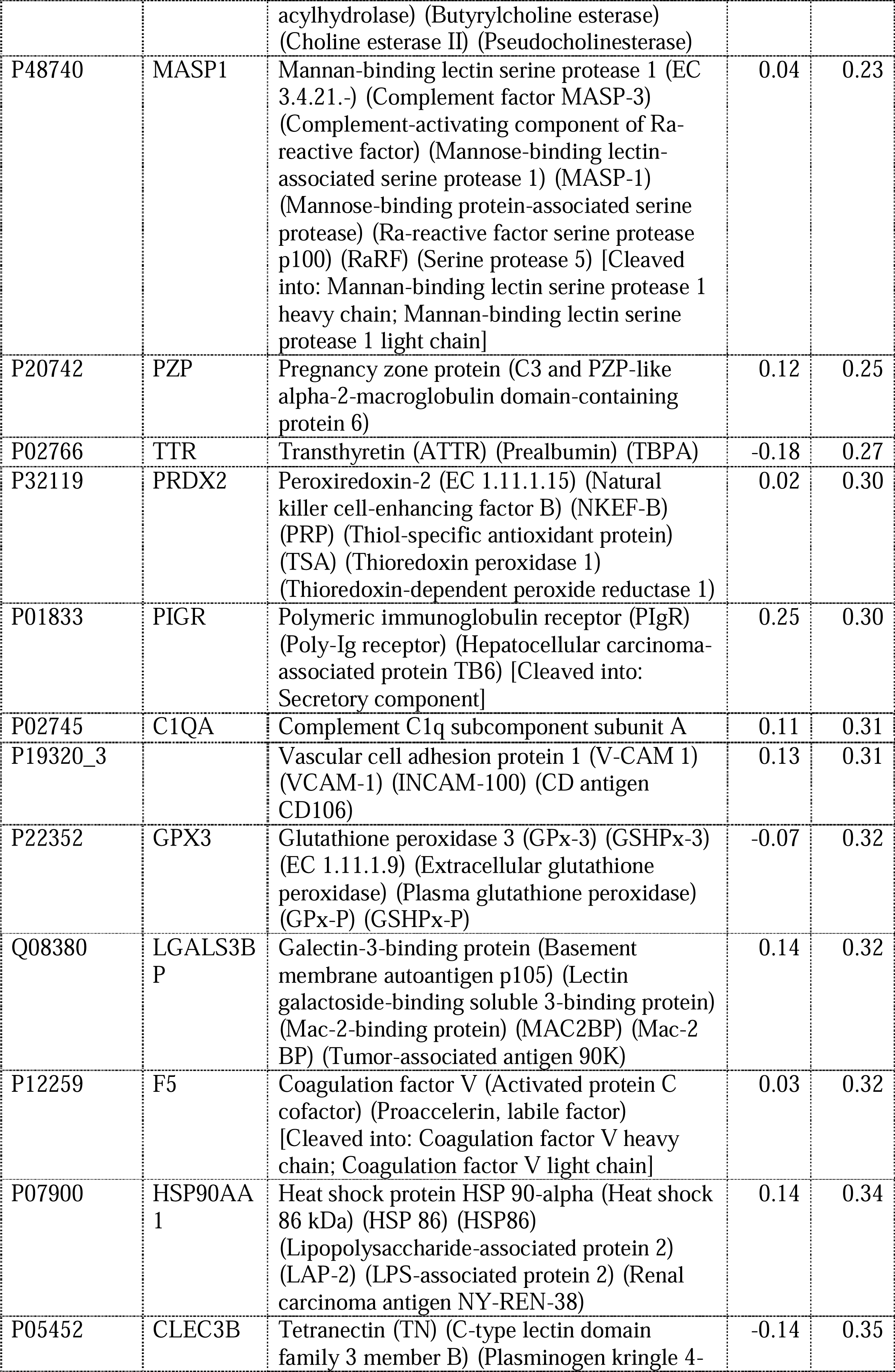
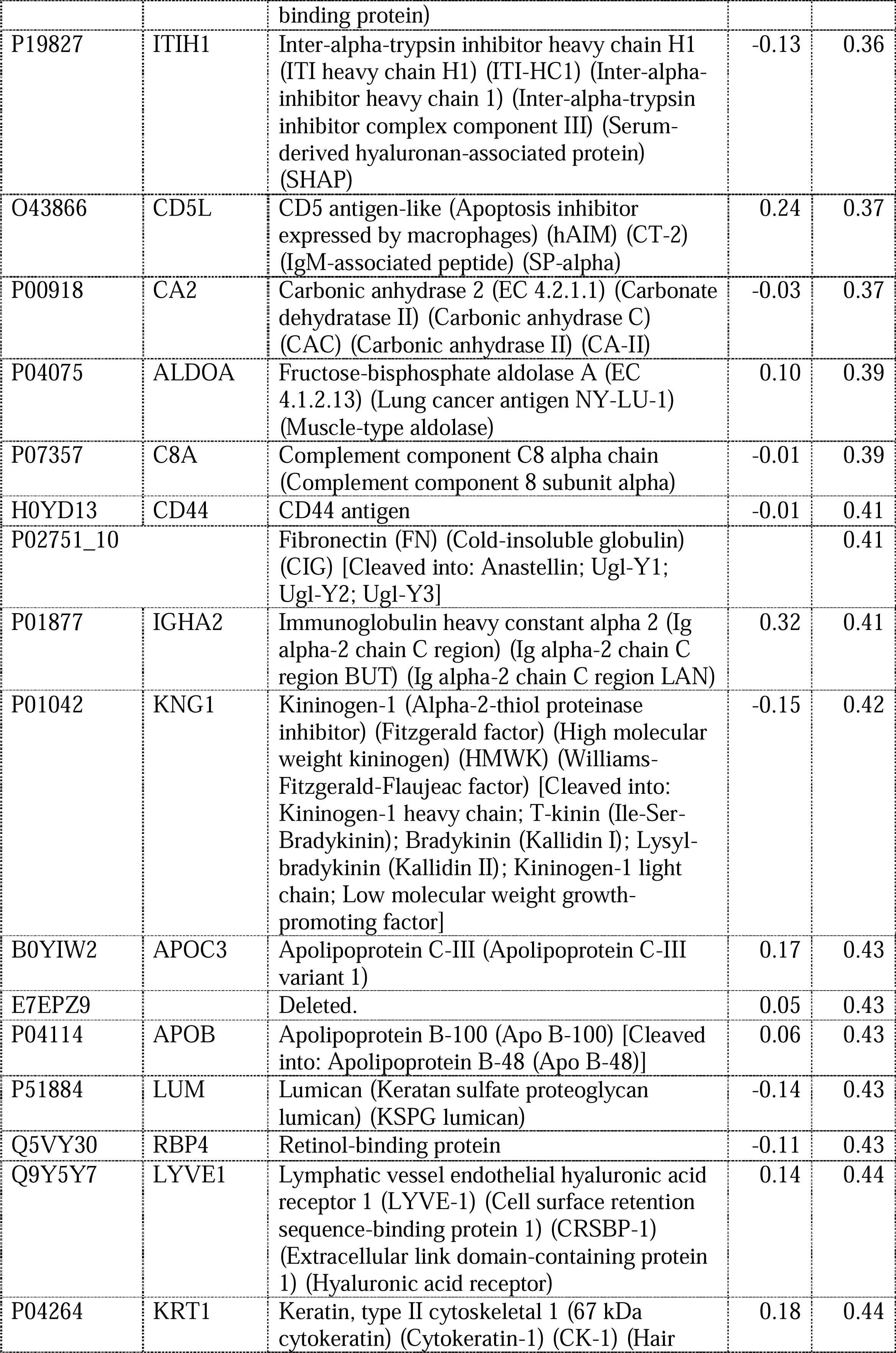
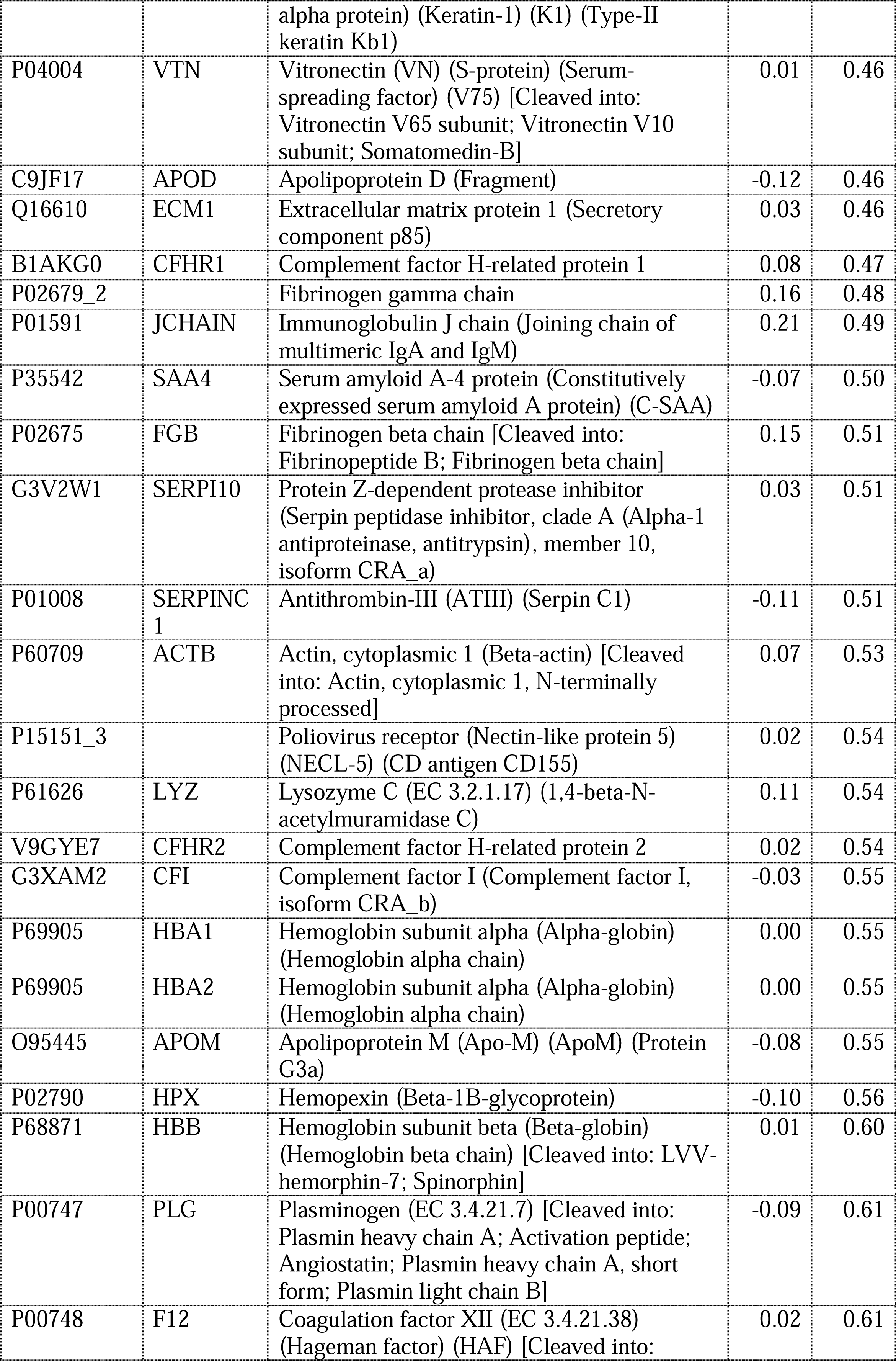
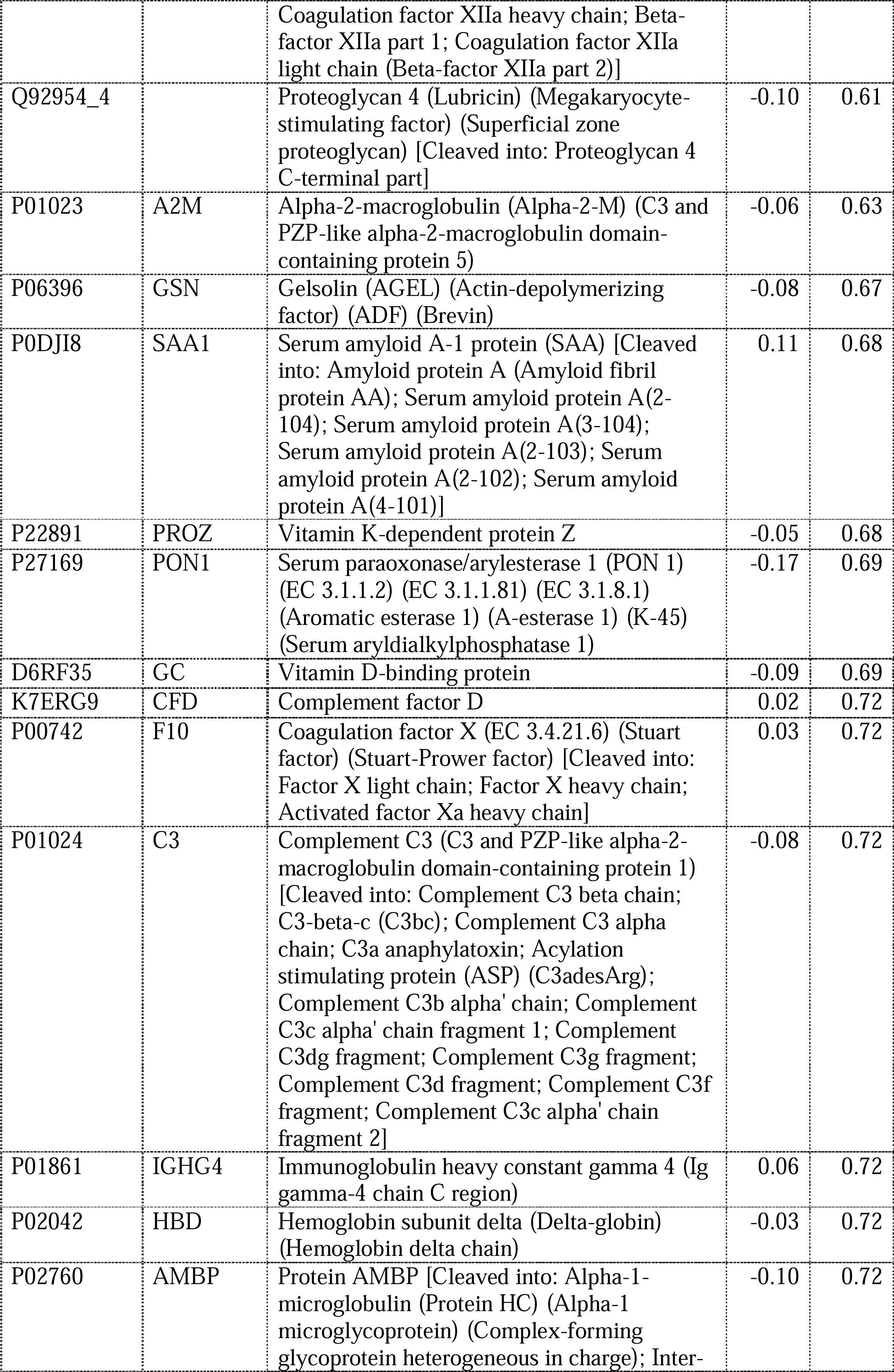
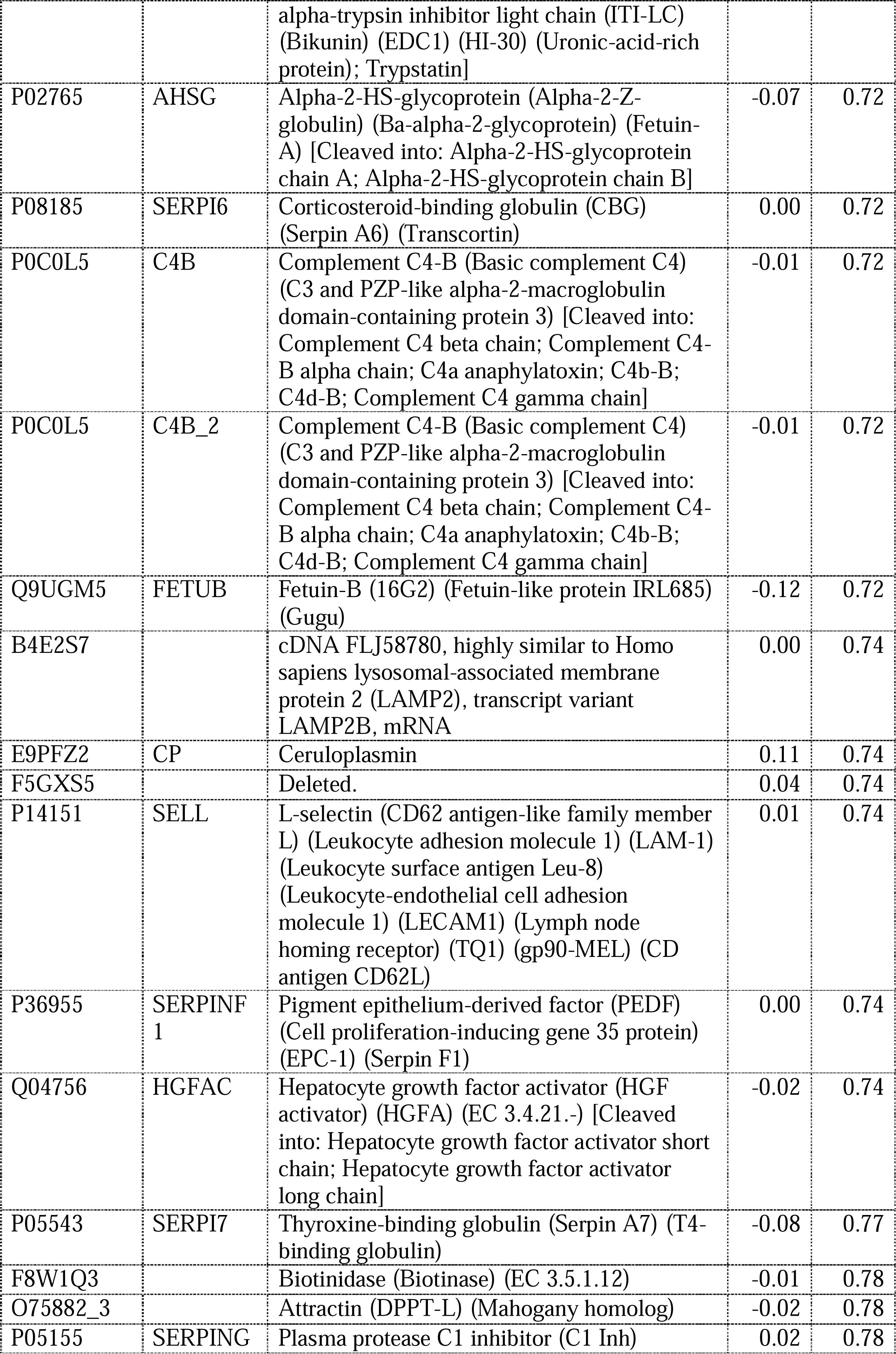
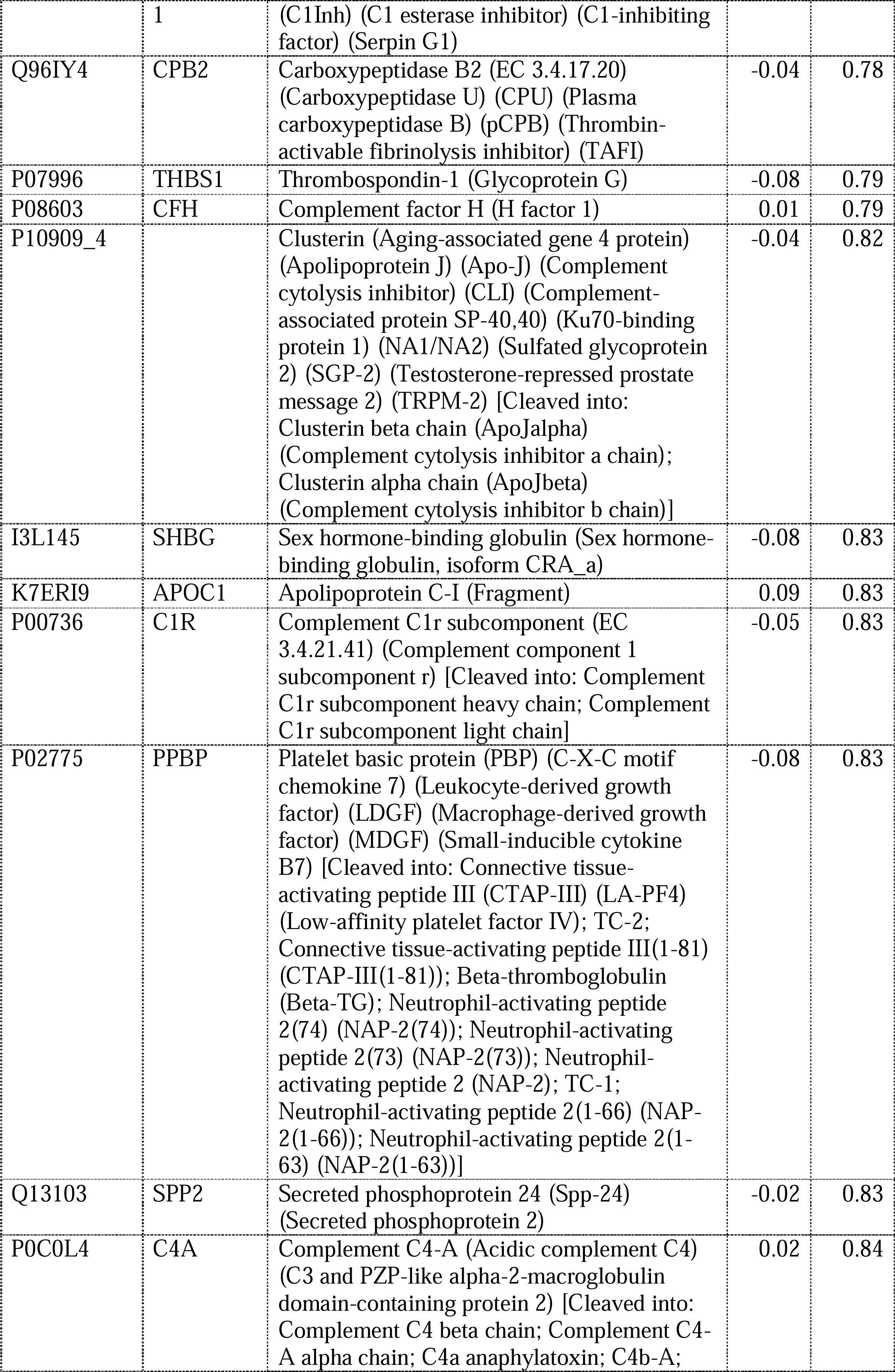
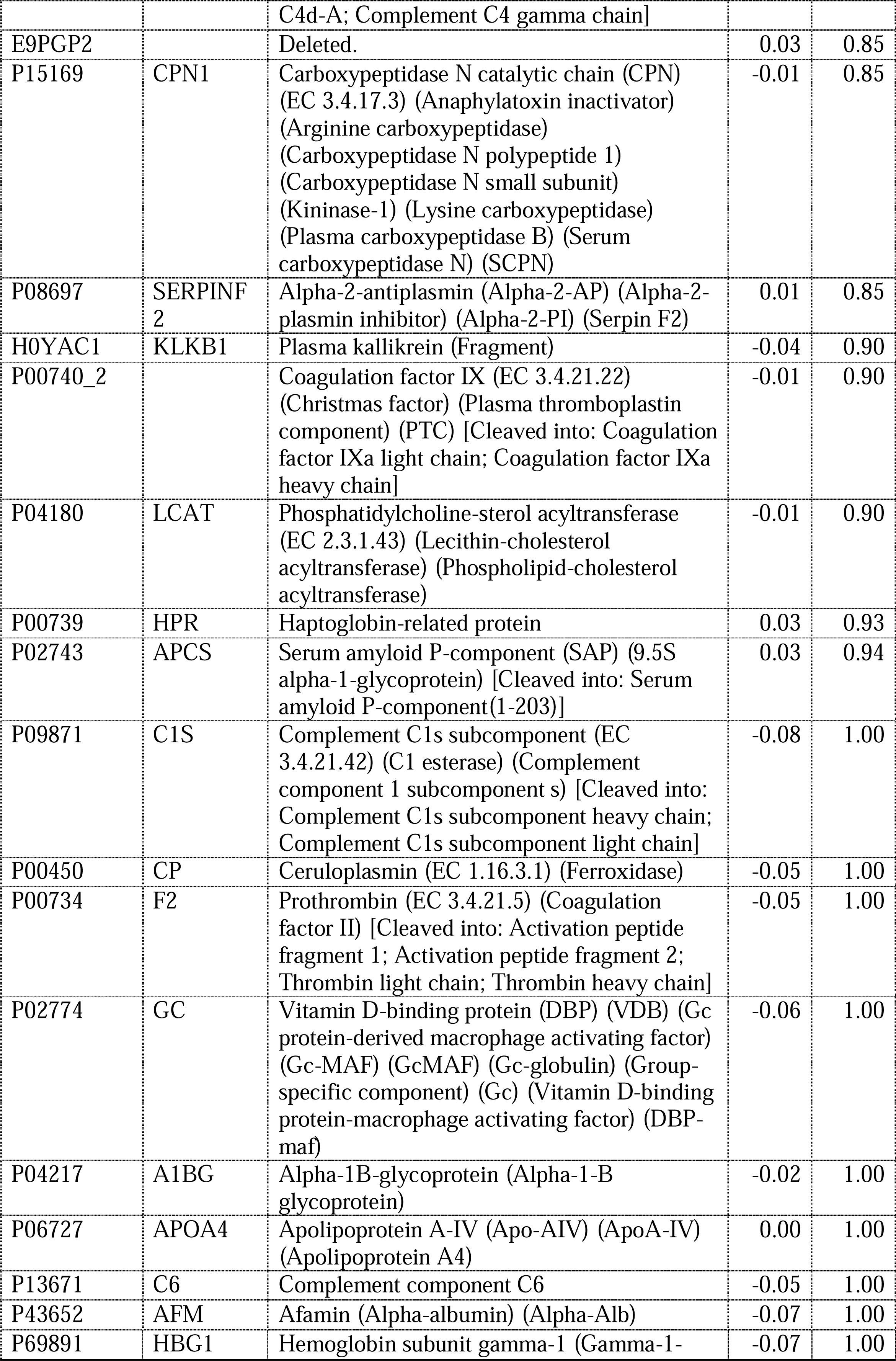
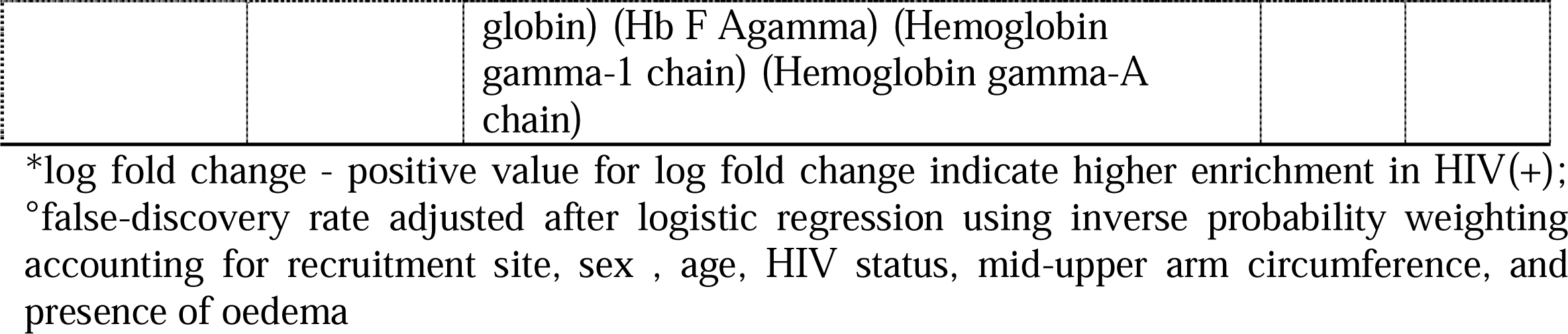
Association between individual proteins to HIV status

